# Circulating Levels of HMGB-1 and NPM/B23 proteins and Clinical Significance in Debut NSCLC patients

**DOI:** 10.1101/2024.11.04.24316686

**Authors:** Tan Hui, Lu Liu, Ying Yi, Carmen Valenzuela, Qiang Zhao, Zhiwei Zhang, Wen Li, Silvio E. Perea, Yasser Perera

**Affiliations:** Cancer Research Institute of Hengyang Medical School, Key Laboratory of Cancer Cellular and Molecular Pathology in Hunan Province, University of South China, 28 Changsheng Road, Hengyang 421001, Hunan Province, China. People’s Republic of China; Department of Pathology, The First Affiliated Hospital of University of South China, Hengyang, 421001, Hunan Province, China. People’s Republic of China; China-Cuba Biotechnology Joint Innovation Center (CCBJIC), Yongzhou Development and Construction Investment Co., Ltd. (YDCI), Yangjiaqiao Street, Lengshuitan District, Yongzhou City, Hunan Province, 425000, China. People’s Republic of China; Institute of Cybernetics, Mathematics and Physics, Havana, Cuba, Calle 15 entre Cy D. Plaza de la Revolución. CP 10400; La Habana, Cuba; Molecular Oncology Group, Department of Pharmaceuticals, Biomedical Research Division, Center for Genetic Engineering & Biotechnology (CIGB), Havana 10600, Cuba

## Abstract

Alarmins are endogenous molecules that alert the immune system to circumvent the invading antigens in the host. They are basically sensed by specific cellular receptors, leading to recruitment and activation of dendritic cells to mount innate and adaptive immune responses. However, a dual role of alarmins as mediators in either antitumor immunity or tumor progression has been also reported. Here, we investigated the status of the HMGB-1 alarmin and the alarmin-like NPM/B23 in 162 debut NSCLC patients and 60 age-matched healthy individuals as control. NSCLC patients exhibited lower median values of circulating HMGB-1 compared to control (30.04 *versus* 37.30 ng/ml, p=0.02), but higher levels of HMGB-1 were positively correlated with tumor size > 2.0 cm (p=0.004) irrespectively of tumor histology. Otherwise, circulating NPM/B23 levels were significantly increased in NSCLC patients respect to control (812 *versus* 551 pg/ml, p=0.00027). Likewise, patients with tumors > 2 cm exhibited higher levels NPM/B23. Strong association was observed between HMGB-1 and NPM/B23 in serum of NSCLC patients (r=0.679 p=0.0001) compared to healthy individuals who exhibited a weaker correlation (r=0.278 p=0.031). However, the strongest correlation between both alarmins was observed in patients with STAS and tumors ≤ 2 cm (r=0.900 p=0.0001). Co-expression of HMGB-1 and NPM/B23 was also observed in tumor tissues although to a lesser extent. Our data unveil that concomitant circulating HMGB-1 and NPM/B23 could serve as a putative alarmin-based biomarker of NSCLC disease progression.

## Introduction

Lung cancer is responsible for a large number of deaths worldwide annually in part due to the late clinical onset of the disease and the scarcity of biomarkers that help to predict early diagnosis, risk of progression, clinical response and survival. However, most of the NSCLC biomarkers so far discovered correspond to actionable gene mutations helping to guide tailored molecular targeted therapies with successful objective clinical response in around 30 % of the patients (1). For this reason, the search for new biomarkers linked to the natural history of the disease in NSCLC patients at onset is a priority in public health worldwide. Besides, such biomarkers should be feasible to measure and preferably by non-invasive methods. The alarmins which have capacity to promote, recruit, activate, and mature dendritic cells (2), could potentially play important roles as soluble biomarkers in diagnosis, cancer progression, and/or treatment of NSCLC. Particularly, in our work, we focused attention on the clinical significance of circulating levels of HMGB-1, which is a widely studied alarm not only in cancer but also in conditions of autoimmunity and diabetes (3). We also explored the coexistence of circulating HMGB-1 with NPM/B23, which has been less studied as alarmin in cancer.

On the other hand, a dual protumor and antitumor behavior has been suggested by other authors (4). For example, intracellular HMGB-1, which is a non-histone chromatin-associated protein, is crucial in tumor growth, influences response to therapy and promotes both cell death and survival (5). However, this alarmin is involved in different pathways associated with cancer progression and can enhance response to immune checkpoint blockade in cancer patients. (5). As putative biomarker in cancer, different evidences have been already reported for HMGB-1. In urothelial carcinoma of bladder elevated levels both in tumor tissue and serum have been found (6); in advanced lung cancer, small cell lung carcinoma (SCLC), and Mesothelioma, high HMGB1 levels did correlate with shorter overall survival in NSCLC patients (7). In pancreatic ductal adenocarcinoma (PDAC), HMGB1 was a better diagnostic biomarker for PDAC compared with pre-existing PDAC biomarkers such as carbohydrate antigen (CA) 19–9 or carcinoembryonic antigen (CEA) CA19–9 and CEA (8), and finally HMGB-1 did predict progression in gastric cancer (9) and response to chemotherapy in breast cancer (10).

On the other hand, intracellular NPM/B23, also known as NPM1, B23, No38, numatrin is a phosphoprotein mainly localized in the nucleolus (11), and it is involved in mRNA transport, chromatin remodeling, genome stability and apoptosis (12). It is typically overexpressed in solid tumors, being associated with mitotic index and metastasis in most cases (13). However, very few data have been reported about the existence of circulating levels of NPM/B23 in the serum of cancer patients and/or a possible role of this as alarms.

To our knowledge, there are no previous studies in which the coexistence of circulating levels of HMGB-1 and NPM/B23 in the serum of cancer patients is of clinical significance in cancer patients. In order to establish groundbreaking evidences, in our study we investigated whether both proteins correlated with each other and/or with clinicopathological parameters of patients with NSCLC debut.

## Patients and Methods

### Subjects

A total of 167 NSCLC patients and 60 age-matched healthy individuals were enrolled in this study. The study was approved by the Ethics Committee of the First Affiliated Hospital of University of South China, Hunan Province and reviewed lot number No.2021LL0115001. Also, the study was conducted in accordance with ethical principles from Helsinki declaration (2013), the regulations on Biomedical Ethics Review involving human (2016), the regulations on Clinical Research Ethics review in traditional Chinese medicine and the International Ethical Guidelines for Biomedical Research. A consent signed by all the subjects included in the study was obtained.

### Enrollment criteria for NSCLC patients and sample collection

Only debut patients without previous chemoradiotherapy, targeted therapy or immunotherapy before surgery, were included. Also, patients with paraffin histology diagnosis after radical resection of lung cancer with LUAD or LUSC were eligible for the study. Those patients with poor quality of serum, other histology types or previous lung cancer surgery were excluded. All the patients’ samples originated pairs of wax blocks (cancer and its corresponding normal lung tissue taken 5 cm away from tumor). Blood samples from patients and healthy individuals were obtained and serum was collected and kept at -80°C until used for measuring HMGB-1 and NPM/B23 by ELISA.

### Measurement of HMGB 1 and NPM/B23 in serum of patients NSCLC

HMGB-1 in serum was quantified used a commercially available Human High-Mobility Protein 1 (HMGB 1) ELISA Kit (ml026429), Shanghai mlbio Technology Co., LTD. On the other hand, NPM/B23 was determined by using a Human Nucleolar Phosphoryl Protein (NPM) ELISA Kit (ml058225), Shanghai mlbio Technology Co., LTD.

### Immunohistochemistry in tissue samples

Paraffin-embedded tissue samples were sliced and submitted to immunohistochemistry analysis by using a Ready to Use Immunohistochemical Hypersensitivity UltraSensitiveTM SP kit (rabbit / rat) (KIT-9729), Fuzhou Maixin Biotechnology Development Co., LTD. Recombinant Anti-HMGB1 Antibody [EPR3507] (ab79823), and Recombinant Anti-Nucleophosmin Antibody [EP1848Y] (ab52644), Abcam (Shanghai) Trading Co., Ltd, were used as primary antibodies for detecting HMGB-1 and NPM/B23 respectively. Data were expressed as positive expression rate which correspond to the number of positive sections respect total sections × 100.

### Statistical Analysis

The quantitative variables are reported as mean, standard deviation (SD), median, interquartile range, and range. The qualitative are reported as absolute and relative frequencies. The expression of NPM/B23 and HMGB-1 in patients and healthy subjects was compared using the Mann-Whitney U test. A generalized linear model was used to explore the association between NPM/B23 and HMGB-1 (each one) regarding clinicopathological parameters. Spearman’s rank correlation was used to assess relationships among NPM/B23 and HMGB-1 globally and stratified by tumoral size and STAS. Statistical analyses were done using SPSS version 26. The level of significance chosen was α = 0.05.

## Results

### Demographic and Clinicopathological Characteristics

Mean age was 61.2 (±8) years in the NSCLC patient group and 62.4 (±11.8) years in the control group. Sex of NSCLC patients was grouped in 98 female (42.2%) and 134 males (57.8%). Thirty male and 30 female subjects were grouped in the control group. The entire NSCLC patient group included 162 subjects (69.8%) diagnosed with LUAD, and 70 subjects (30,2%) diagnosed with LSCC. Clinical stages were grouped in two major cohorts: stage I and stages II, III, IV. The mean tumor size was 2.3 (±1.3) cm according to the major diameter and the frequency of other clinical conditions like presence or absence of lymphatic metastasis (23 *versus* 191 subjects), pleural invasion (23 *versus* 191 subjects), and STAS (79 *versus* 135 subjects) were also taken into account in our study. The degree of differentiation was classified as Low (65 subjects), Mid (125 subjects), Mid-Low (5 subjects), High (23 subjects) or High-Mid (1 subject) (Table 1). Patients with miss information or useless samples were not eligible for correlation analysis. Clinical outcome to standard anticancer therapy and overall survival of patients are being registered at the moment.

**Table 1.** Association between circulating HMGB1 levels and the clinicopathological characteristics.

|  |  | Cohorts |  |  |  |
| --- | --- | --- | --- | --- | --- |
|  |  | Patients |  |  | Healthy |
|  |  | N(162) | HMGB-1 (ng/ml) | N(60) | HMGB-1 (ng/ml) |
| Gender | Female | 74 | 29849.4 (19033.2) | 30 | 37572.6 (9935.8) |
|  | Male | 88 | 30242.1 (20416.8) | 30 | 35879.8 (11216.1) |
|  | Mann-Whitney-U |  | 0.872 |  | 0.525 |
| Age | ≤60 | 76 | 32308.2 (23580.2) | 29 | 32556.1 (11260.8) |
|  | >60 | 86 | 28642.2 (16532.3) | 31 | 37597.3 (6227.3) |
|  | Mann-Whitney-U |  | 0.092 |  | 0.110 |
| Smoking history | No | 120 | 29873.4 (19110.4) |  | -- |
|  | Yes | 42 | 30725.8 (24519.4) |  |  |
|  | Mann-Whitney-U |  | 0.582 |  |  |
| Histological type | LUAD | 124 | 29640.9 (19260.5) |  | -- |
|  | LUSC | 38 | 34180.4 (27807.9) |  |  |
|  | Mann-Whitney-U |  | 0.083 |  |  |
| Degree of differentiation | Low | 38 | 29841.5 (22337.8) |  | -- |
|  | Mid | 90 | 29997.5 (18852.9) |  |  |
|  | High | 21 | 31382.4 (17229.9) |  |  |
|  | Mann-Whitney-U |  | 0.837 |  |  |
| Clinical Stage | I | 126 | 29873.4 (20013.1) |  | -- |
|  | II-III-IV | 36 | 30622.6 (18760.6) |  |  |
|  | Mann-Whitney-U |  | 0.581 |  |  |
| Lymphatic metastasis | No | 133 | 29942.9 (20158.7) |  | -- |
|  | Yes | 11 | 30036.4 (7519.1) |  |  |
|  | Mann-Whitney-U |  | 0.848 |  |  |
| Pleural invasion | No | 128 | 29873.4 (20064.3) |  | -- |
|  | Yes | 16 | 30448.5 (11728.8) |  |  |
|  | Mann-Whitney-U |  | 0.995 |  |  |
| STAS | No | 95 | 29803.7 (16108.1) |  | -- |
|  | Yes | 49 | 30052.1 (28006.6) |  |  |
|  | Mann-Whitney-U |  | 0.454 |  |  |
| Tumor size (major diameter) | ≤ 2 | 96 | 28520.1 (13880.2) |  | -- |
|  | > 2 | 48 | 33051.5 (30920.6) |  |  |
|  | Mann-Whitney-U |  | **0.004\*** |  |  |

### Levels of Circulating Alarmins in NSCLC patients

As shown in Figure 1A, median values of HMGB-1 levels were significantly lower in NSCLC patients (30.04 ng/ml) compared to healthy individuals (37.30 ng/ml) (p=0.02). Interestingly, LUAD but not LUSC patients exhibited significant reduction of the HMGB-1 levels compared to healthy individuals (p<0.01). However, HMGB-1 levels in LUAD and LUSC patients were not significantly different between both groups. Otherwise, the median values of B23-Nucleophosmin levels were significantly increased in NSCLC patients (812 pg/ml) compared with healthy individuals (551 pg/ml) (p=0.001) (Fig. 1B). When both histological types were analyzed separately, the circulating B23-Nucleophosmin levels were also significantly increased in LUAD and LUSC patients respect to control group (p<0.001) (Fig. 1B). No significant difference of circulating B23-Nucleophosmin levels between LUAD and LUSC patients were observed.

**Figure 1:**
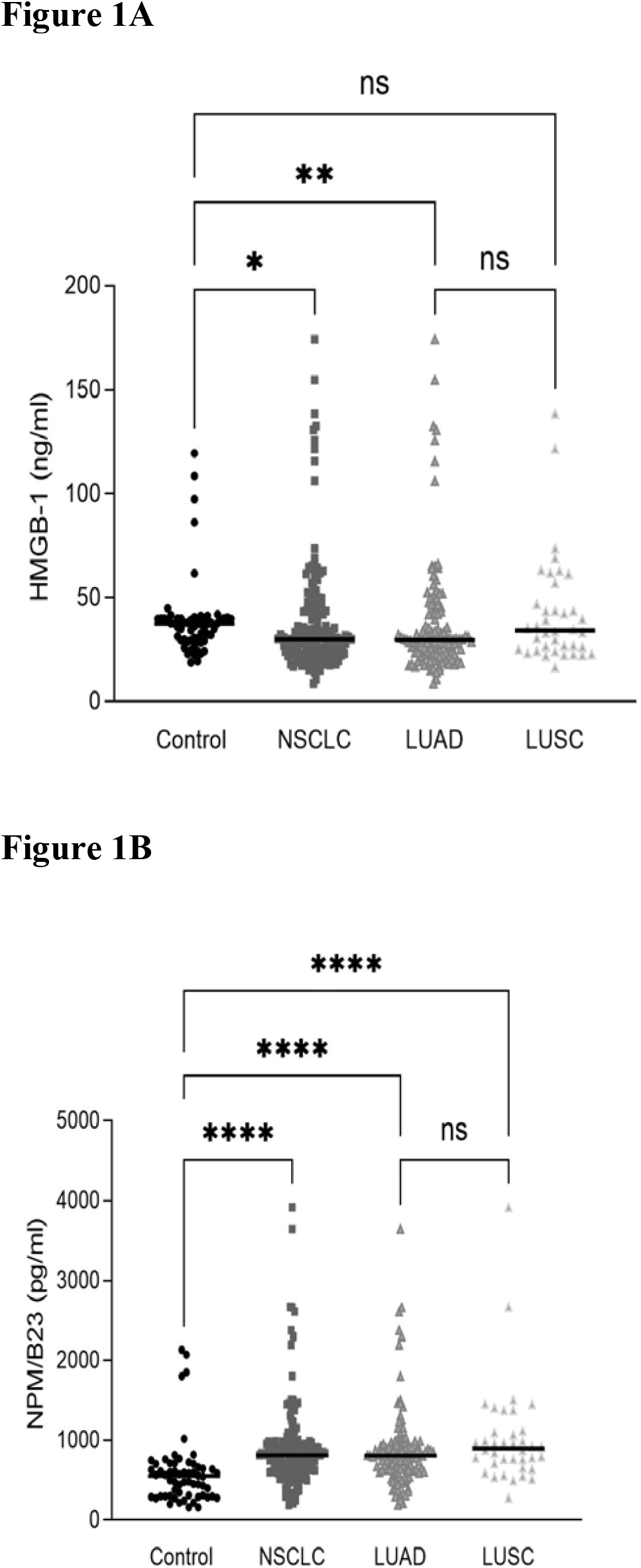
Levels of circulating alarmins were determined by ELISA in serum from 162 NSCLC patients and 60 healthy individuals as Control. NSCLC patients were also analyzed according to LUAD and LUSC histology. A: HMGB-1 and B: NPM/B23.

### Association between circulating alarmins levels and clinicopathological features of NSCLC patients

Once the levels of both alarmins in the serum of the patients were investigated, we next interrogated whether such levels were associated to any specific clinicopathological feature in NSCLC patients. Interestingly, data shown in Table 1 exhibit a strong association between tumor size and the HMGB-1 levels in serum of NSCLC as determined by univariate analysis. Particularly, significant higher levels of HMGB-1 were positively correlated with tumor size > 2.0 cm according to major diameters of lesions (p=0.004) and irrespectively of tumor histology. Correlation between circulating HMGB-1 and other clinicopathological features were not observed according to this analysis. When generalized linear models were performed using HMGB1 as dependent variable and considering all the features gathered, only tumor size was also confirmed to be associated (Table S1, Supplementary-1).

We also looked for putative association between circulating NPM/B23 levels with specific clinicopathological features in NSCLC patients. Data shown in Table 2 indicate that presence of STAS in NSCLC patients and tumor size > 2 cm were associated with higher levels of NPM/B23 in serum with p=0.043 and p=0.0001 respectively. However, data from multivariate analysis shown that only tumor size > 2 cm was strongly associated with the circulating NPM/B23 levels (Table S2, Supplementary-1).

**Table 2.**
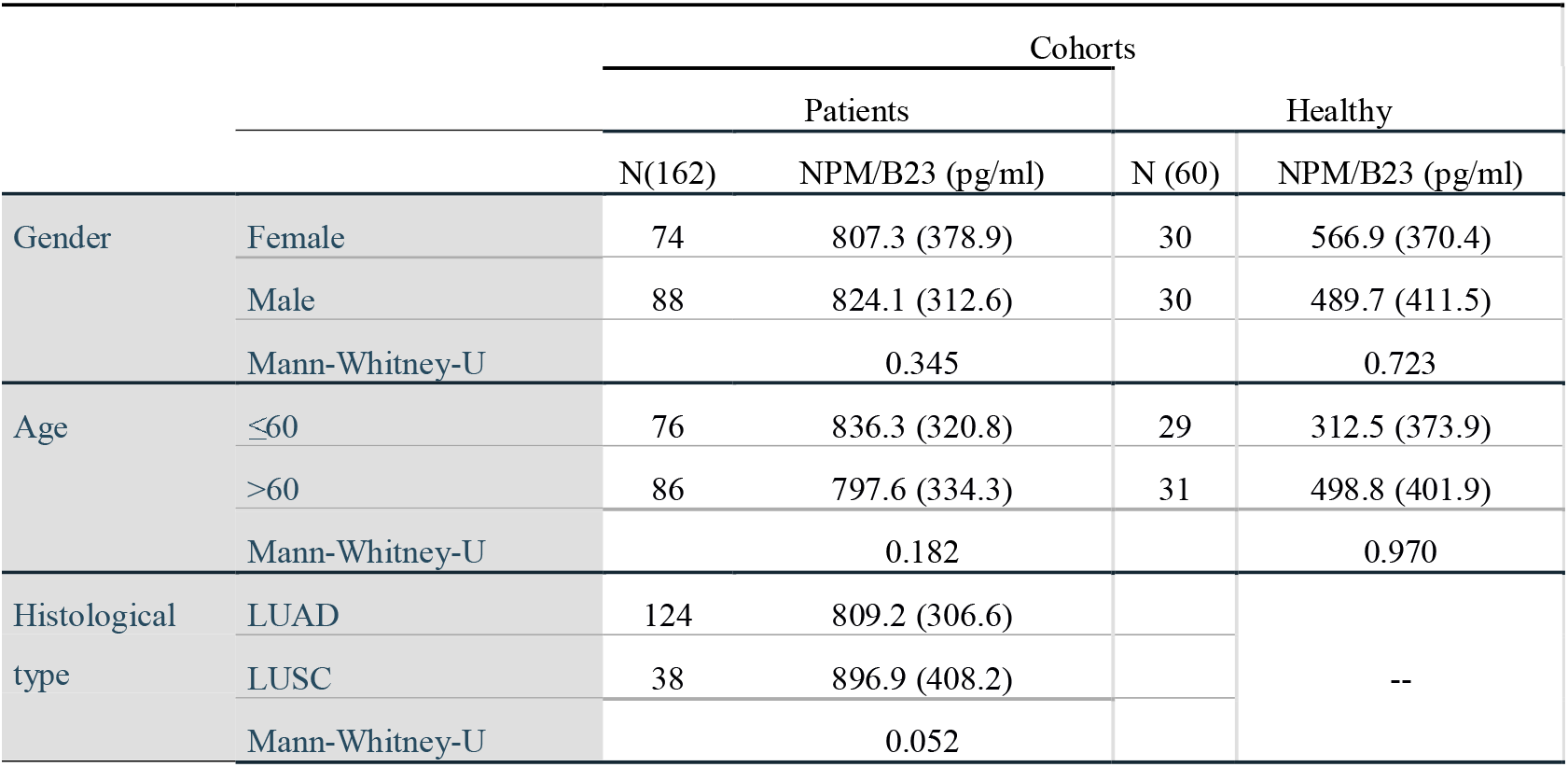
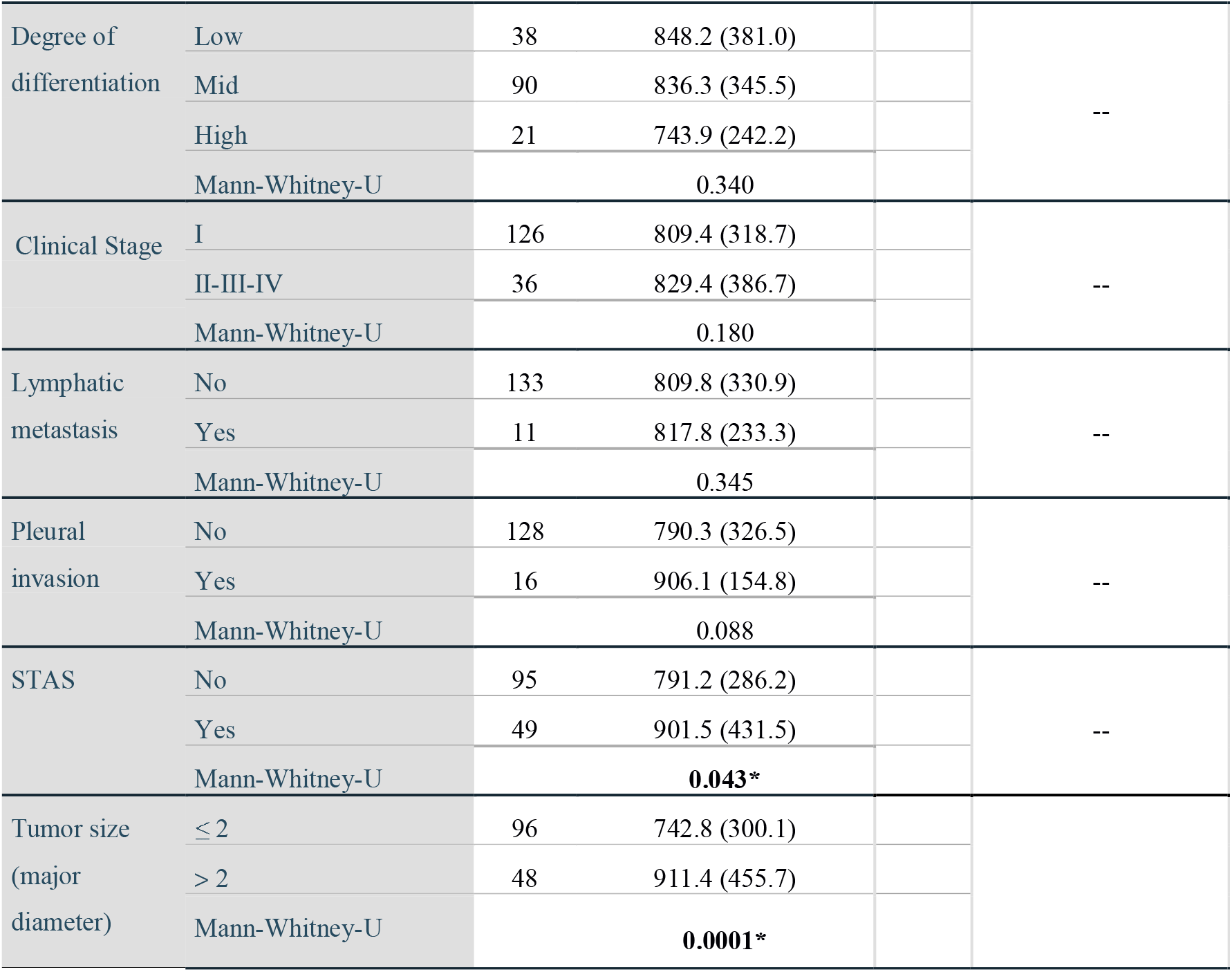
Association between circulating NPM/B23 levels in serum and the clinicopathological features.

To verify whether circulating HMGB-1 and NPM/B23 levels did correlate each other, we analyzed both alarmins in serum from healthy individuals and in NSCLC patients. Data in Table 3 indicated that levels of HMGB-1 and NPM/B23 were correlated in both cohorts, however such correlation was stronger in NSCLC patients compared to healthy individuals (r=0.679 *versus* r=0.278).

**Table 3.** Correlation between HMGB-1 and NPM/B23 levels in serum.

| Table 3. Correlation between HMGB-1 and NPM/B23 levels in serum |  |  |  |  |  |
| --- | --- | --- | --- | --- | --- |
| Cohorts |  |  |  | HMGB-1 | NPM/B23 |
| Spearman's rho | Healthy | HMGB1 (ELISA) | Correlation Coefficient | 1.000 | .278* |
|  |  |  | Sig. (2-tailed) | . | .031 |
|  |  |  | N | 60 | 60 |
|  | NSCLC Patients | HMGB1 (ELISA) | Correlation Coefficient | 1.000 | .679** |
|  |  |  | Sig. (2-tailed) | . | .0001 |
|  |  |  | N | 162 | 162 |

\*, Correlation is significant at the 0.05 level (2-tailed).
\*\*, Correlation is significant at the 0.01 level (2-tailed).

As tumor size and presence of STAS in NSCLC patients seemed to be associated with the circulating alarmin levels in our study, we subsequently investigated the correlation between HMGB-1 and NPM/B23 levels in these two selected clinicopathological features. Interestingly, in patients with tumors > 2 cm the correlation between both alarmins was significant (near r=0.70) irrespectively of STAS (Table 4). However, the strongest correlation between both circulating alarmins was observed in NSCLC patients with tumors ≤ 2cm and presence of STAS (r=0.900, p=0.0001) (Table 4).

**Table 4.**
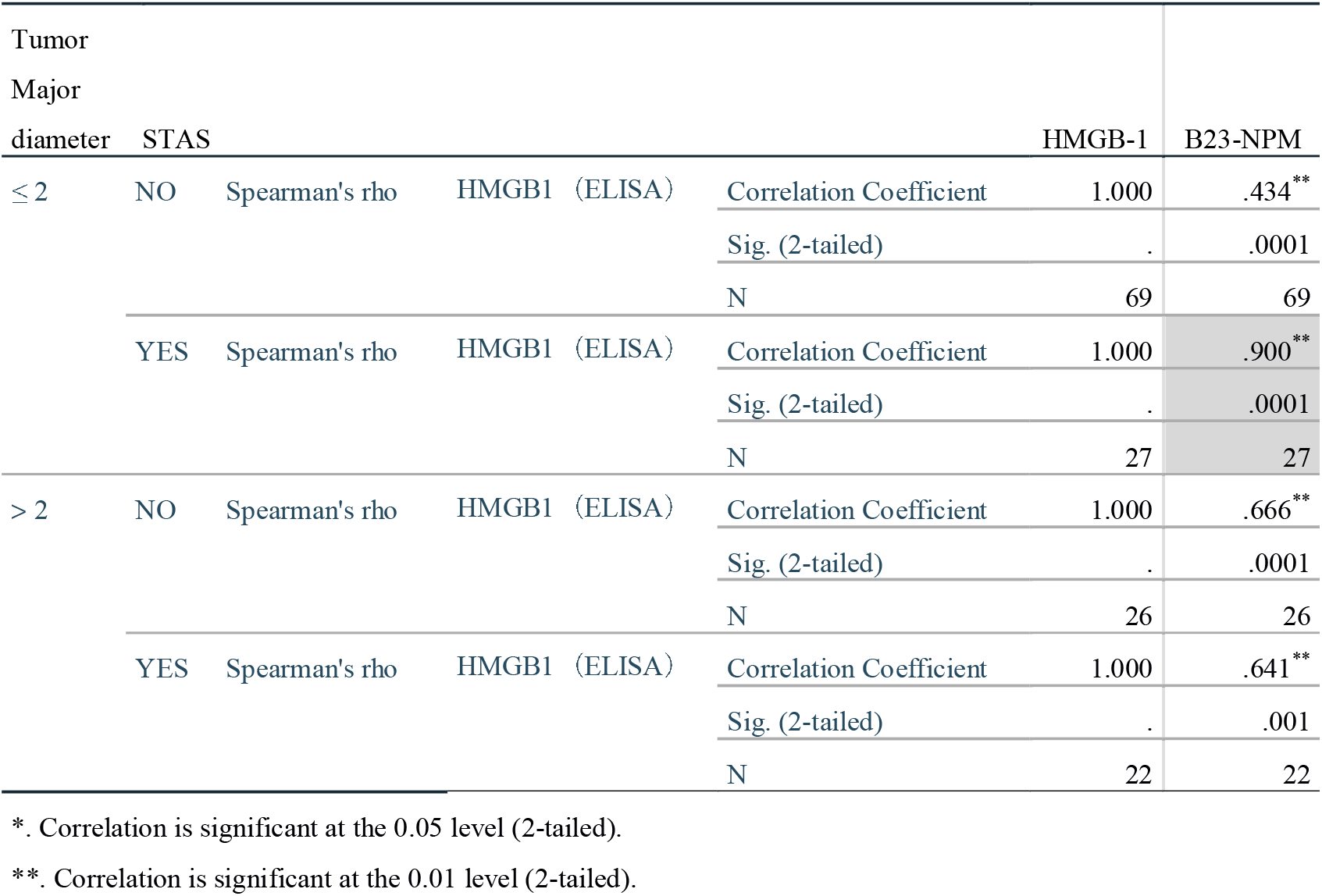
Correlation between HMBG1 and NPM/B23 according to tumor size and presence of STAS.

Finally, we wanted to explore the intratumor expression of both alarmins in NSCLC patients by immunohistochemistry in tumor biopsies and adjacent healthy tissues 5 cm away from tumor as internal comparator. Data in Supplem. 2 show illustrative examples of intracellular HMGB-1 expression both in tumor tissues and adjacent healthy tissues by immunohistochemistry. This alarmin was mainly expressed in the nuclear compartment and it was not only confined to alveolar epithelial cells but also phagocytes in the alveolar space, fiber cells in the alveolar septum, and fibrocytes.

As immunohistochemistry analysis is rather semiquantitative, the HMGB-1 positive expression rate was considered as the sum of patients exhibited low (+), moderate (++) and high (+++) intracellular levels. Of note, the results in Table 5 showed that among 167 NSCLC patients, the positive rate of HMGB-1 expression in cancer tissues was 78.44%, while in adjacent healthy lung tissues was 86.83% (p=0.043). Even more interesting was a dichotomic HMGB-1 expression pattern observed for both NSCLC histology types included in this study. In LUAD patients the positive expression rate of HMGB-1 protein was lower than that in adjacent healthy lung tissue (p=0.0001); otherwise in LUSC diagnosed patients the positive rate was higher in tumor tissues compared to adjacent healthy tissues (p=0.001). Nonetheless, it is interesting that more than 50% of positive expression rate was detectable in adjacent healthy tissues from NSCLC patients. We next looked for potential correlation between intracellular HMGB-1 levels in tumors and adjacent healthy tissues. A significant although weak positive correlation was observed between intracellular levels of this alarmin in tumors and in their normal tissue counterpart (Table S3, Supplementary-1) in NSCLC patients (p=0.0001; r=0.348). However, no correlation was observed between circulating HMGB-1 levels in sera and tumor or normal tissues (Table S3).

**Table 5.** Positive expression rate of HMGB-1 in NSCLC patients by immunohistochemistry.

|  |  |  |  |  |  |  |  |  |
| --- | --- | --- | --- | --- | --- | --- | --- | --- |
| Tissue Type | Patients | HMGB1 expression level |  |  |  | Positive rate | X<sup>2</sup> | p value |
|  |  | (-) | (+) | (++) | (+++) |  |  |  |
| Tumor NSCLC | 167 | 36 | 12 | 49 | 70 | 78.44% | 4.089 | 0.043 |
| Healthy adjacent | 167 | 22 | 14 | 39 | 92 | 86.83% |  |  |
| Tumor LUAD | 130 | 31 | 9 | 38 | 52 | 76.92% | 24.071 | 0.0001 |
| Healthy adjacent | 130 | 4 | 6 | 30 | 90 | 96.92% |  |  |
| Tumor LUSC | 37 | 5 | 3 | 11 | 18 | 83.78% | 10.660 | 0.001 |
| Healthy adjacent | 37 | 18 | 8 | 9 | 2 | 51.35% |  |  |

On the other hand, immunohistochemistry analysis shown a nuclear or nuclear/nucleolar expression pattern for the NPM/B23 alarmin (Supp 3). Contrary to what observed for HMGB-1, the overall positive rate of NPM/B23 on tumor tissues was higher than that observed for adjacent healthy tissues (92.22% *versus* 85.03%; p=0.0387) (Table 6). Likewise, the positive expression rate in tumor tissues from LUAD patients was 91.54 % *versus* 83.08% observed for adjacent healthy tissues. Interestingly, in LUSC patients the positive expression rate of NPM/B23 was not significantly different between tumor and adjacent healthy tissues (94.60% *versus* 91.89%; p=0.6433) (Table 6). Nonetheless, the NPM/B23 positive expression rate was very high in NSCLC patients. In line with HMGB-1, a significant although weak positive correlation was also observed between intracellular levels of NPM/B23 in tumors and in their normal tissue counterpart (Table S4, Supplementary-1) in NSCLC patients (p=0.0001; r=0.394). Likewise, no correlation was observed between circulating NPM/B23 levels in sera and tumor or normal tissues (Table S4).

**Table 6.** Positive expression rate of B23/NPM in NSCLC patients by immunohistochemistry.

|  |  |  |  |  |  |  |  |  |
| --- | --- | --- | --- | --- | --- | --- | --- | --- |
| Tissue Type | Patients | NPM/B23 | | | | Positive rate | $\chi^2$ | p value |
|  |  | (-) | (+) | (++) | (+++) |  |  |  |
| Tumor NSCLC | 167 | 13 | 4 | 15 | 135 | 92.22% | 4.276 | 0.0387* |
| Healthy adjacent | 167 | 25 | 47 | 87 | 8 | 85.03% |  |  |
| Tumor LUAD | 130 | 11 | 4 | 9 | 106 | 91.54% | 4.200 | 0.0404* |
| Healthy adjacent | 130 | 22 | 43 | 57 | 8 | 83.08% |  |  |
| Tumor LUSC | 37 | 2 | 0 | 6 | 29 | 94.60% | 0.215 | 0.6433 |
| Healthy adjacent | 37 | 3 | 4 | 30 | 0 | 91.89% |  |  |

As both HMGB-1 and NPM/B23 exhibited high positive expression rate in adjacent healthy tissues in NSCLC patients, we additionally assessed the Ki-67 protein which is a cell proliferation biomarker and is not an alarmin to verify specificity. As expected, a very weak expression rate of Ki-67 in adjacent healthy tissues was only observed compared to tumor tissues in NSCLC patients (2.2% *versus* 56.7% (p<0.001).

To verify whether HMGB-1 and NPM/B23 alarmins also correlate in tissues, the spearman coefficient analysis was analyzed in malignant and normal tissues. Unlike to what we observed in serum, only weak correlation between both proteins was observed in tumor tissues but not in healthy tissues from NSCLC patients (p=0.02; r=0.223) (Table S5/S6, Supplementary-1).

## Discussion

Early detection of new disease biomarkers in NSCLC not only can optimize clinical decision making on therapy but also might supplement CT-scan and chest radiography to mitigate cancer-related mortality and improve overall prognosis in lung cancer. As above-mentioned, the alarmins have been suggested as biomarkers in diagnosis, cancer progression, and/or treatment of NSCLC. In fact, previous meta-analysis studies have indicated that the HMGB-1 alarmin is found to be overexpressed in tumors and serum of NSCLC patients compared to healthy individuals (16) and a potential biomarker to predict progression and survival of NSCLC (17)(18). However, other results demonstrating lower levels of HMGB1 expression in NSCLC patients’ tissue compared to matched normal lung tissues have also been reported (19). Here, we investigated the relevance of circulating and tissue pretherapeutic levels of the HMGB-1 alarmin and also of MPN/B23, which had been suggested as a secreted alarmin in sepsis and found elevated in NSCLC patients (14)(15). In particular, we investigated the clinical significance of positivity for both proteins in relation to the essential clinicopathological parameters of NSCLC patients at the time of onset, as well as the correlation between HMGB-1 and NPM/B23 at both the systemic and tissue levels in tumors.

Our results demonstrated that median global circulating HMGB-1 values in the serum of NSCLC patients were not higher than those observed in healthy individuals but rather tended to be lower, particularly in patients diagnosed with LUAD (Fig. 1A). It is known today that different oxidation states of HMGB-1 leads to three mutually exclusive functions: alarmin (20), chemoattraction (21) or tolerance (22). Therefore, it would be interesting to know whether a specific HMGB-1 redox status is taking place in LUAD patients which could be linked to clinical outcome, progression disease, or survival. Nonetheless, our results showed that elevated individual HMGB-1 levels were positively associated with tumor size > 2 cm (p=0.004)(Table 1) irrespective of the histological type of NSCLC. This finding may be relevant to the clinical staging of patients and indicative of tumor progression for both LUAD and LUSC. These results are consistent with previous ones that show a positive correlation of circulating HMGB-1 levels with tumor size, among other clinicopathological features (17). Although we did not find a correlation of HMGB-1 with other clinicopathological parameters of NSCLC patients, our findings are the first to setup a tumor size threshold of 2 cm defining the significance of the correlation between HMGB-1 and this clinical parameter. Concerning the histology type, we cannot rule out that the non-discrimination between LUAD and LUSC in our study could be due to the group imbalance between both histological types (124 versus 38, respectively).

NPM/B23 is a multifunctional protein relevant in cancer with chaperone activity and in ribosomal biogenesis and its intracellular overexpression has been reported in cancer patients (11)(12)(13). However, little evidence has been published about extracellular NPM/B23 as well as their possible role as alarmins. Other authors reported elevated circulating levels of NPM/B23 in patients with NSCLC compared to healthy individuals, a positive correlation with the presence of nodules, and the histological type LUAD showed higher levels of NPM/B23 compared to LUSC (15). In line with these previous results and contrary to what was observed for HMGB-1, in our study we found significantly higher levels of NPN/B23 in NSCLC patients of both the LUAD and LUSC histological types (Fig. 1B). Even more interestingly, as observed for HMGB-1, in the uni- and multivariate analysis there was an association between elevated levels of NPM/B23 and tumor size > 2 cm (p=0.0001) (Table 2), irrespective of the histological type. Additionally, the presence of STAS was correlated with high circulating levels of NPM/B23, although with a weaker significance (p=0.043). Because the presence of STAS in NSCLC is indicative of early spread of tumor cells through the airspaces adjacent to the primary tumor (23), any biomarker associated with this clinical condition can in principle provide valuable prognostic information and aid in the stratification of the risk of progression. Univariate and multivariate analyses did not show interactions of interest between NPM/B23 and other clinicopathological features. In other words, both HMGB-1 and NPM/B23 individually were associated with clinical parameters indicative of disease progression, such as tumor size and the presence of STAS, regardless of the histological type of NSCLC. In the case of B23/NPM, our findings in debut patients along with those previously reported (15) contribute to pave the way for a possible role of NPM/B23 as an alarmin in NSCLC.

Once the clinical relevance of HMGB-1 and NPM/B23 had been analyzed separately, we proceeded to evaluate the correlation between both proteins in serum and tissues. Importantly, our results demonstrated a positive correlation between both proteins in the serum of NSCLC patients at debut (Table 3), which helps to reinforce the hypothesis that NPM/B23 could also behave as an alarmin in this clinical niche. Considering that the individual analysis of circulating HMGB-1 and NPM/B23 were associated with parameters of clinical disease progression such as tumor size and the presence of STAS, we wanted to know if this correlation was maintained according to such clinical-pathological parameters. Importantly, a strong positive correlation was found between HMGB-1 and NPM/B23 when tumors were smaller than 2 cm in diameter and in the presence of STAS (p=0.0001; r=0.900) (Table 4). For tumors larger than 2 cm, a weak and significant correlation was observed between both alarmins irrespectively of the presence of STAS. To our knowledge, our findings are the first to report a direct positive correlation between circulating levels of HGMB-1 and NPM/B23 in cancer patients as well as their association with a range of tumor size and early tumor cell spreading. Although elevated individual levels of circulating HMGB1 and NPM/B23 may reveal changes associated with disease progression, the strong correlation observed between both proteins in smaller tumors accompanied by STAS may be useful for early detection of the disease and/or prediction of the risk of NSCLC progression in conjunction with non-invasive imaging procedures such as ct-scan and Magnetic Resonance Images. In addition, early detection may help to oncologist practitioners in determining more valuable therapeutic strategies. For this reason, our results deserve to be validated in a larger patient population.

Taking into account that alarmins can be released from tumor cells as part of the immune response that is generated during the tumor-host interaction as well as necrotic processes inherent to solid tumors, we evaluated the levels of HMGB-1 and NPM/B23 expression in tumor tissue and in healthy lung tissue adjacent to the tumor. Our findings indicated that although the HMGB-1 expression positivity rate in NSCLC patient tumors was greater than 50%, the frequency of HMGB-1 expression positivity in tumor tissue was lower than that observed in matched adjacent healthy tissues, especially in LUAD patients (Table 5). However, in LUSC patients, the behavior was rather reversed, perhaps as expected according to the alarming function of HMGB-1. Further studies are necessary to know what is the real significance of the differential expression of HMGB-1 in tumor tissue versus adjacent healthy lung tissue in LUAD patients (Table 6). On the other hand, the positive expression rate of NPM/B23 was greater than 80% in both tumor tissue and healthy tissue, which is a novel aspect in our work. In contrast, the expression of Ki-67, which is a biomarker of cell proliferation and not an alarmin, behaved as expected with a very low positive expression rate (2.2%) in the healthy tumor tissue of NSCLC patients. In order to know if the co-expression of HMGB-1 and NPM/B23 in the tumor behaved similarly to that observed with the circulating levels of both proteins, the correlation analysis was carried out. Interestingly, a weak correlation (p=0.02; r=0.223) was observed, which should be strengthened in future studies with a larger number of patients with NSCLC. No relevant associations of both intracellular proteins with individual clinicopathological parameters were observed.

All in all, in this work we have investigated the individual and correlative status of HMGB-1 and NPM/B23 alarmins in serum and tissues of NSCLC patients who have not been treated with any anticancer therapeutic option. Our data are the first in describing correlations between both proteins in serum of NSCLC patients in association with clinical status at the onset of the disease.

## Conclusions

Our findings describe a strong correlation between circulating levels of HMGB-1 and NPM/B23 with clinical significance in terms of early disease progression of NSCLC. Tumor size of 2 cm was found to be a threshold defining such strong correlation. Both, circulating HMGB-1 and NPM/B23 were also associated with tumor size irrespective histology. Positive co-expression of HMGB-1 and NPM/B23 in tumor tissue tended to be correlative as well, although a larger number of patients is needed to strength the data.

## Supporting information

Supplemental Figure IHC of HMGB-1

Supplemental Figure IHC of NPM/B23

## Data Availability

All data produced in the present work are contained in the manuscript

## Conflict declaration

Nothing to declare

## Financial support

This work was supported by MOST “National key R&D program of China (2021YFE0192100)”

## Notes

### Competing Interest Statement

The authors have declared no competing interest.

### Author Declarations

The study was approved by the Ethics Committee of the First Affiliated Hospital of University of South China, Hunan Province and reviewed lot number No.2021LL0115001. Also, the study was conducted in accordance with ethical principles from Helsinki declaration (2013), the regulations on Biomedical Ethics Review involving human (2016), the regulations on Clinical Research Ethics review in traditional Chinese medicine and the International Ethical Guidelines for Biomedical Research. A consent signed by all the subjects included in the study was obtained.

## References

1. Kathryn CA, and Gregory JR. SystemicTherapy for Locally Advanced and Metastatic Non–Small Cell Lung Cancer. A Review. JAMA 2019, 322:764–774.

2. Yingjie N, De Yang, and Oppenheim JJ. Alarmins and Antitumor Immunity. Clinical Therapeutics 2016, 38:1042–1053.

3. Jing Z, Longmin C, Faxi W, Yuan Z, Jingyi L, Jiahui L, Faheem K, Fei S, Yang L, Jing L, Zhishui C, Shu Z, Fei X, Qilin Y, Jinxiu L, Kun H, Bao-Ling A, Zhiguang Z, Decio LE, Ping Y, Cong-Yi W. Extracellular HMGB1 exacerbates autoimmune progression and recurrence of type 1 diabetes by impairing regulatory T cell stability. Diabetologia 2020 19:987–1001. doi: 10.1007/s00125-020-05105-8

4. Damien B and Eicke L. HMGB1, IL-1α, IL-33 and S100 proteins: dual-function alarmins. Cellular & Molecular Immunology 2017, 14:43–64.

5. Sourour I, Takwa B, Shona P, Mohamed E, Izzaldin A, Sabah A, Said D, Maysaloun M, Shahab U. Role of HMGB1 and its associated signaling pathways in human malignancies. Cellular Signalling 2023, 112:110904. Doi:10.1016/j.cellsig.2023.110904

6. Singh A, Gupta N, Khandakar H, Kaushal S, Seth A, Pandey RM, Sharma A. Autophagy-associated HMGB-1 as a novel potential circulating non-invasive diagnostic marker for detection of Urothelial Carcinoma of Bladder. Mol. Cell Biochem 2022, 477:493–505.

7. Xia Q, Xu J, Chen H, Gao Y, Gong F, Hu L, Li Y. Association between an elevated level of HMGB1 and non-small-cell lung cancer: a meta-analysis and literature review. OncoTargets and Therapy 2016, 9:3917–3923.

8. Chung HW, Lim JB, Jang S, Lee KJ, Park KH, Song SY. Serum high mobility group box-1 is a powerful diagnostic and prognostic biomarker for pancreatic ductal adenocarcinoma. Cancer Sci 2012, 103:1714–1721.

9. Chung H, Lee SG, Kim H, Hong D, Chung J, Stroncek D, Lim J-B. Serum high mobility group box-1 (HMGB1) is closely associated with the clinical and pathologic features of gastric cancer. J. Transl. Med 2009, 7:38 doi: 10.1186/1479-5876-7-38.

10. Stoetzer OJ, Fersching DM, Salat C, Steinkohl O, Gabka CG, Hamann U, Braun M, Feller A-M, Heinemann V, Siegele B, Nagel D, Holdenrieder S. Circulating immunogenic cell death biomarkers HMGB1 and RAGE in breast cancer patients during neoadjuvant chemotherapy. Tumour Biol 2013, 34:81–90.

11. Grisendi S, Mecucci C, Falini B, Pandolfi PP. Nucleophosmin and cancer. Nat Rev Cancer 2006; 6: 493–505.

12. Mikael SL. NPM1/B23: A Multifunctional Chaperone in Ribosome Biogenesis and Chromatin Remodeling. Biochem Res Int. 2010, 2011:195209. doi: 10.1155/2011/195209

13. Siying C, Hairong H, Yan W, Leichao L, Yang L, Haisheng Y, Yalin D, Jun L. Poor prognosis of nucleophosmin overexpression in solid tumors: a meta-analysis. BMC Cancer 2018,18:838. doi: 10.1186/s12885-018-4718-6.

14. Nawa Y, Kawahara K-i, Tancharoen S, Meng X, Sameshima H, Ito T, Masuda Y, Imaizumi H, Hashiguchi T, and Maruyama I. Nucleophosmin may act as an alarmin: implications for severe sepsis. Journal of Leukocyte Biology 2009, 86:645–53.

15. Akın G, Esbah O, Eröz R. Could Nucleolin and Nucleophosmin Levels Be Prognostic Indicators in Non-Small Cell Lung Cancer? Acta facultatis medicae Naissensis 2022; 39: 433–442.

16. Xia Q, Xu J, Chen H, Gao Y, Gong F, Hu L and Yang L. Association between an elevated level of HMGB1 and non-small-cell lung cancer: a meta-analysis and literature review. OncoTargets and Therapy 2016, 9:3917–3923.

17. Feng A, Tu Z, and Yin B. The effect of HMGB1 on the clinicopathological and prognostic features of non-small cell lung cancer. Oncotarget 2016, 7:20507–20519

18. Handke NA, Aba R, Trimpop N, von Pawel J, Holdenrieder S. Soluble high mobility group box 1 (HMGB1) is a promising biomarker for prediction of therapy response and prognosis in advanced lung cancer patients. Diagnostics 2021, 11: 356 doi: 10.3390/diagnostics11020356

19. Shen X, Hong L, Sun H, Shi M, Song Y. The expression of high-mobility group protein box 1 correlates with the progression of non-small cell lung cancer. Oncol Rep 2009:535–539.

20. Yang H, Lundbäck P, Ottosson L, Erlandsson-Harris H, Vénéreau E, Bianchi ME, Al-Abed Y, Andersson U, Tracey KJ, Antoine DJ. Redox modification of cysteine residues regulates the cytokine activity of high mobility group box-1 (HMGB1). Mol Med 2012, 18:250–259.

21. Vénéreau E, Casalgrandi M, Schiraldi M, Antoine DJ, Cattaneo A, De Marchis F, Liu J, Antonelli A, Preti A, Raeli L, Shams SS, Yang H, Varani L, Andersson U, Tracey KJ, Bachi A, Uguccioni M, Bianchi ME. Mutually exclusive redox forms of HMGB1 promote cell recruitment or proinflammatory cytokine release. J Exp Med 2012, 209:1519–1528.

22. Kazama H, Ricci J-E, Herndon JM, Hoppe G, Green DR, Ferguson TA. Induction of immunological tolerance by apoptotic cells requires caspase-dependent oxidation of high-mobility group box-1 protein. Immunity 2008, 29:21–32.

23. Jia M, Yu S, Gao H, Sun P-L. Spread Through Air Spaces (STAS) in Lung Cancer: A Multiple-Perspective and Update Review. Cancer Manag Res 2020, 23:2743–2752.

