## Supplemental Tables for "Circulating Levels of HMGB-1 and NPM/B23 proteins and Clinical Significance in Debut NSCLC patients"

**Supplementary-1**

**Table S1**. Generalized Linear Models using HMGB1 as dependent variable

| Parameter | B | Std. Error | 95% Wald Confidence Interval | | | Hypothesis Test | | |
| --- | --- | --- | --- | --- | --- | --- | --- | --- |
|  |  |  | Lower | Upper | Wald Chi-Square | | df | Sig. |
| (Intercept) | 40464.5 | 10921.2 | 19059.4 | 61869.7 | 13.7 | | 1 | .000 |
| [Gender=Female] | 447.0 | 5781.5 | -10884.6 | 11778.6 | 0.0 | | 1 | .938 |
| [Age≤60] | 1564.2 | 4523.3 | -7301.4 | 10429.8 | 0.1 | | 1 | .729 |
| [Smoking history=NO ] | 5459.3 | 6628.2 | -7531.8 | 18450.4 | 0.7 | | 1 | .410 |
| [Diff_fusion=High ] | 3549.4 | 6928.9 | -10031.0 | 17129.9 | 0.3 | | 1 | .608 |
| [Diff_fusion=Low ] | -1677.1 | 5920.8 | -13281.7 | 9927.5 | 0.1 | | 1 | .777 |
| [lymphatic metastasis=NO ] | 15003.9 | 13716.4 | -11879.9 | 41887.6 | 1.2 | | 1 | .274 |
| [Pleural invasion=NO ] | 179.9 | 8980.7 | -17422.0 | 17781.7 | 0.0 | | 1 | .984 |
| [STAS=NO ] | -8693.4 | 5256.6 | -18996.1 | 1609.4 | 2.7 | | 1 | .098 |
| [Clinical stages=1] | -9219.8 | 11582.7 | -31921.5 | 13482.0 | 0.6 | | 1 | .463 |
| [Histology=LUSC ] | 2817.1 | 8061.3 | -12982.7 | 18617.0 | 0.1 | | 1 | .426 |
| [Tumor size ≤ 2] | -11761.0 | 5438.3 | -22419.9 | -1102.2 | 4.7 | | 1 | .031 |

**Table S2.** Generalized Linear Models using B23 as dependent variable

| Parameter | B | Std. Error | 95% Wald Confidence Interval | | | Hypothesis Test | | |
| --- | --- | --- | --- | --- | --- | --- | --- | --- |
|  |  |  | Lower | Upper | Wald Chi-Square | | df | Sig. |
| (Intercept) | 1241.1 | 216.0 | 817.7 | 1664.5 | 33.0 | | 1 | .000 |
| [Gender=Female] | -8.4 | 114.4 | -232.5 | 215.7 | 0.0 | | 1 | .941 |
| [Age≤60] | 11.0 | 89.5 | -164.4 | 186.3 | 0.0 | | 1 | .902 |
| [Smoking history=NO ] | 90.8 | 131.1 | -166.2 | 347.7 | 0.5 | | 1 | .489 |
| [Diff_fusion=High ] | -39.4 | 137.1 | -308.1 | 229.2 | 0.1 | | 1 | .774 |
| [Diff_fusion=Low ] | -72.4 | 117.1 | -301.9 | 157.1 | 0.4 | | 1 | .536 |
| [lymphatic metastasis=NO ] | 129.0 | 271.3 | -402.7 | 660.8 | 0.2 | | 1 | .634 |
| [Pleural invasion=NO ] | -108.5 | 177.6 | -456.6 | 239.7 | 0.4 | | 1 | .541 |
| [STAS=NO ] | -182.2 | 104.0 | -386.0 | 21.6 | 3.1 | | 1 | .080 |
| [Clinical stages=1] | -161.3 | 229.1 | -610.3 | 287.7 | 0.5 | | 1 | .481 |
| [Histology=LUSC ] | 146.0 | 159.4 | -166.5 | 458.6 | 0.8 | | 1 | .360 |
| [Tumor size ≤ 2] | -235.5 | 107.6 | -446.3 | -24.7 | 4.8 | | 1 | .029 |

**Table S3: Spearman Coefficient analysis of HMGB-1 in serum, tumor and healthy tissues**

|  | | | HMGB1（Serum）ELISA | HMGB1(Cancer) IHC |
| --- | --- | --- | --- | --- |
| Spearman's rho | HMGB1（Serum）ELISA | Correlation Coefficient | 1.000 | -.028 |
|  |  | Sig. (2-tailed) | . | .742 |
|  |  | N | 162 | 136 |
|  | HMGB1(Cancer) IHC | Correlation Coefficient | -.028 | 1.000 |
|  |  | Sig. (2-tailed) | .742 | . |
|  |  | N | 136 | 188 |
|  | HMGB1（Healthy tissues） IHC | Correlation Coefficient | -.147 | .348^**^ |
|  |  | Sig. (2-tailed) | .087 | .0001 |
|  |  | N | 136 | 188 |

**Table S4: Spearman Coefficient analysis of NPM/B23 in serum, tumor and healthy tissues**

|  | | | NPM/B23  Serum (ELISA) | NPM/B23  (Cancer) IHC |
| --- | --- | --- | --- | --- |
| Spearman's rho | NPM/B23(Serum) ELISA | Correlation Coefficient | 1.000 | -.064 |
|  |  | Sig. (2-tailed) | . | .454 |
|  |  | N | 162 | 138 |
|  | NPM/B23(Cancer) IHC | Correlation Coefficient | -.064 | 1.000 |
|  |  | Sig. (2-tailed) | .454 | . |
|  |  | N | 138 | 190 |
|  | NPM/B23（Healthy tissues） IHC | Correlation Coefficient | -.127 | .394^**^ |
|  |  | Sig. (2-tailed) | .138 | .0001 |
|  |  | N | 138 | 190 |

**Table S5: Spearman Coefficient analysis between HMGB-1 and NPM/B23 in tumor tissues.**

|  | | | NPM/B23 (Cancer) IHC | HMGB-1 (Cancer) IHC |
| --- | --- | --- | --- | --- |
| Spearman's rho | NPM/B23 (Cancer) IHC | Correlation Coefficient | 1.000 | .223^**^ |
|  |  | Sig. (2-tailed) | . | .002 |
|  |  | N | 190 | 188 |
|  | HMGB-1(Cancer) IHC | Correlation Coefficient | .223^**^ | 1.000 |
|  |  | Sig. (2-tailed) | .002 | . |
|  |  | N | 188 | 188 |
| **. Correlation is significant at the 0.01 level (2-tailed). | | | | |

| **Table S6: Spearman Coefficient analysis between HMGB-1 and NPM/B23 in healthy tissues.** | | | | |
| --- | --- | --- | --- | --- |
|  | | | NPM/B23（Healthy tissues） IHC | HMGB1（Healthy tissues） IHC |
| Spearman's rho | B23-NPM（Healthy tissues） IHC | Correlation Coefficient | 1.000 | .137 |
|  |  | Sig. (2-tailed) | . | .060 |
|  |  | N | 190 | 188 |
|  | HMGB1（Healthy tissues） IHC | Correlation Coefficient | .137 | 1.000 |
|  |  | Sig. (2-tailed) | .060 | . |
|  |  | N | 188 | 188 |
