## Supplemental Figure IHC of HMGB-1 for "Circulating Levels of HMGB-1 and NPM/B23 proteins and Clinical Significance in Debut NSCLC patients"

### Slide 1
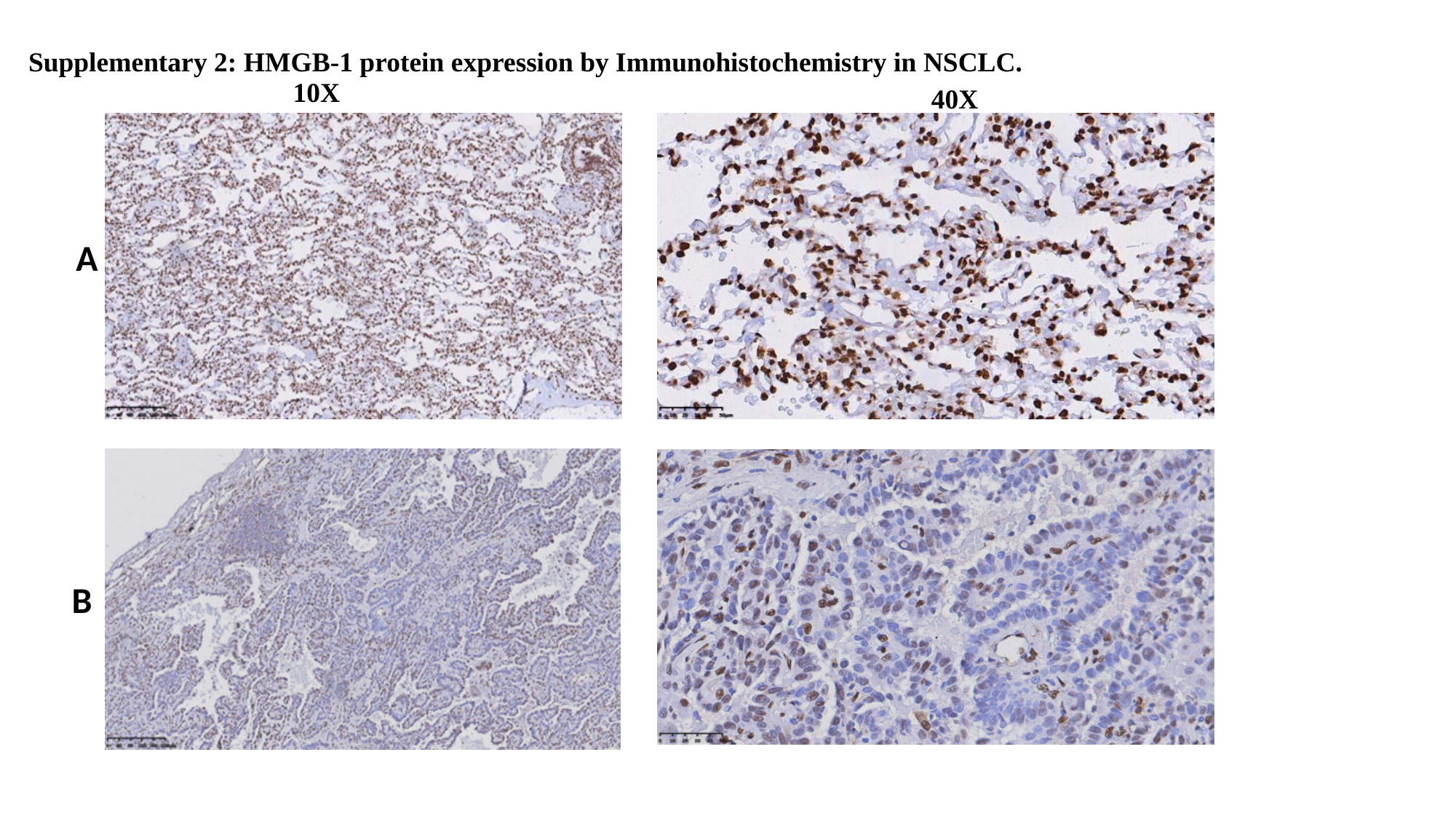

#### Supplementary 2: HMGB-1 protein expression by Immunohistochemistry in NSCLC.
10X
40X
A
B

### Slide 2
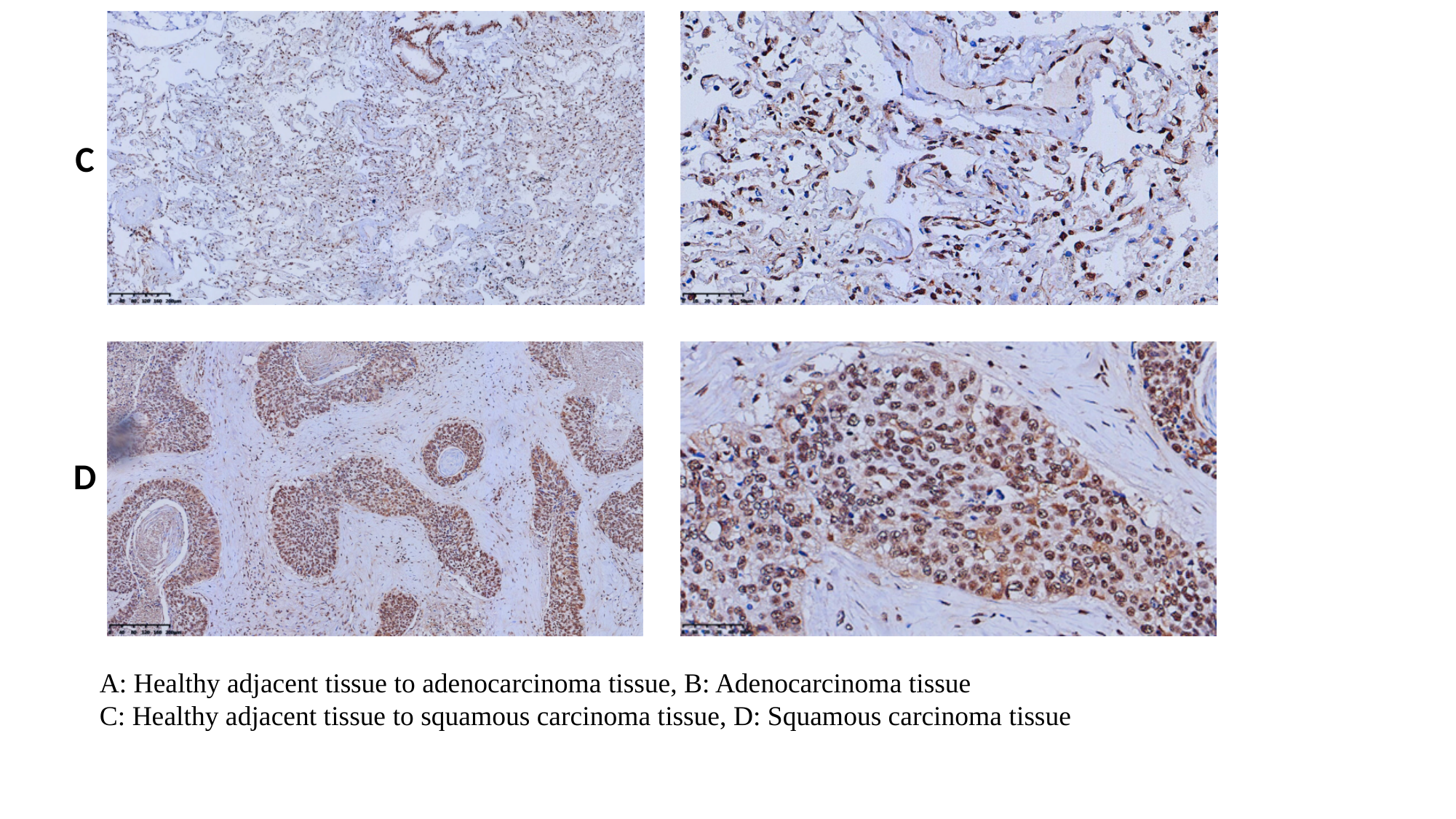

C
D
A: Healthy adjacent tissue to adenocarcinoma tissue, B: Adenocarcinoma tissue
C: Healthy adjacent tissue to squamous carcinoma tissue, D: Squamous carcinoma tissue
